# RAY: CRISPR diagnostic for rapid and accurate detection of SARS-CoV2 variants on a paper strip

**DOI:** 10.1101/2021.02.01.21250900

**Authors:** Manoj Kumar, Sneha Gulati, Asgar Hussain Ansari, Rhythm Phutela, Sundaram Acharya, Poorti Kathpalia, Akshay Kanakan, Ranjeet Maurya, Janani Srinivasa Vasudevan, Aparna Murali, Rajesh Pandey, Souvik Maiti, Debojyoti Chakraborty

## Abstract

The COVID-19 pandemic originating in the Wuhan province of China in late 2019 has impacted global health, causing increased mortality among elderly patients and individuals with comorbid conditions. During the passage of the virus through affected populations, it has undergone mutations- some of which have recently been linked with increased viral load and prognostic complexities. Interestingly, several of these variants are point mutations that are difficult to diagnose using the gold standard quantitative real-time PCR (qPCR) method. This necessitates widespread sequencing which is expensive, has long turn-around times, and requires high viral load for calling mutations accurately. In this study, we show that the high specificity of *Francisella novicida* Cas9 (FnCas9) to point mismatches can be successfully adapted for the simultaneous detection of SARS-CoV2 infection as well as for detecting point mutations in the sequence of the virus obtained from patient samples. We report the detection of the mutation N501Y (earlier shown to be present in the British N501Y.V1, South African N501Y.V2, and Brazilian N501Y.V3 variants of SARS-CoV2) within an hour using paper strip chemistry. The results were corroborated using deep sequencing. Our design principle can be rapidly adapted for other mutations, highlighting the advantages of quick optimization and roll-out of CRISPR diagnostics (CRISPRDx) for disease surveillance even beyond COVID-19.

## INTRODUCTION

Testing and isolating SARS-CoV2 positive cases has been one of the main strategies for controlling the rise and spread of the coronavirus pandemic that has affected about one hundred million people all over the world (WHO). Like all other viruses, the SARS-CoV2 genome has undergone several mutations during its passage through the human host, and the exact positive or negative impacts of most of which are still under investigation^1-3^.

Since December 2020, health authorities in the United Kingdom have reported the emergence of a new SARS-CoV2 variant (lineage B.1.1.7) that is phylogenetically distinct from other circulating strains in the region. With 23 mutations, this variant had rapidly replaced other SARS-CoV2 lineages occurring in the region and has been established to have greater transmissibility through modelling and clinical correlation studies^4-7^. More recently, South African and Brazilian authorities have also reported variants (B.1.351 and P.1, respectively) that have also been associated with a higher viral load in preliminary studies^8-10^. As air travel has resumed in most countries post lockdown, infected individuals harboring these mutations have instantly been detected in countries far away from their origin, generating concerns about greater disease incidence unless these cases are recognized and isolated. This is particularly important as vaccine roll-out has been initiated in several countries, and the studies about vaccine efficacy against the new variants are only in their infancy^11^.

Among the diagnostic methods that have been employed for identifying positive coronavirus cases, qPCR has been the most widely adopted nucleic acid-based test due to its ability to identify even low copy numbers of viral RNA in patient samples and has been considered as a gold standard in diagnosis^12-14^. However, probe-based qRT PCR assays rely on amplification of a target gene with real-time analysis of DNA copy numbers and are not suited for genotyping point mutations associated with novel CoV2 variants^15-18^. Although strategies such as Amplification Refractory Mutation System-quantitative PCR (ARMS-qPCR) have been described in the literature for genotyping mutations in nucleic acids, these require substantial design and validation for reproducible readouts and hence are not routinely used for clinical diagnosis^19,20^. Similarly, antigen-based SARS CoV2 assays which have lower sensitivity than qPCR have not yet been reported to specifically identify mutant variants. In the absence of any true diagnostic test for the variants, most countries have resorted to next-generation sequencing of patient samples-which greatly increases both the time and cost for identifying positive individuals^21-24^.

We and several others have documented the applicability of CRISPR diagnostics (CRISPRDx) for identifying CoV2 signatures in patient samples^25-36^. Using the Cas9 from *Francisella novicida* (FnCas9) we have recently reported the development of the FELUDA (FnCas9 Editor Linked Uniform Detection Assay) platform for SARS-CoV2 diagnosis with similar accuracy as the gold standard qPCR test^25^. In this report, we introduce RAY (Rapid variant AssaY), an advancement of the FELUDA platform to identify mutational signatures of the coronavirus variants in a sample eliminating the need for sequencing. RAY can successfully detect both the infection as well as the presence of the common N501Y mutation present across all the three new variants and distinguish it from the parent CoV2 lineage. Thus it can be employed as a primary surveillance method for isolating cases belonging to these groups and can enable the initiation of appropriate management protocols.

## RESULTS

### Identification of mutations in SARS-CoV2 variants for CRISPR targeting

To develop RAY for SARS-CoV2 variant identification, we first analyzed the mutations arising in the three variant lineages reported from the UK, South Africa, and Brazil and looked for the accessibility to FnCas9 enzyme based on the presence of an NGG PAM site in the vicinity^37^. In an earlier study, we had successfully established that FnCas9 is unable to bind or cleave targets having two mismatches -in the 2nd and 6th position (PAM proximal) of the sgRNA-with respect to the target (Figure 1A)^25,38^. Besides these, we had also systematically identified other regions in the sgRNA where mismatches lead to weak or no substrate binding. We found that out of the 32 defining SNVs reported across all the three lineages^37^, 10 sites were targetable by FnCas9 (Figure 1B, C, Supplementary Table 1). Critically, the N501Y mutation present across all the lineages falls within this category^39^. This mutation is present in the receptor-binding domain of the virus and is the subject of numerous studies analyzing the efficacy of vaccines against the mutated form of the virus^40-42^. Owing to its prevalence as a shared marker for mutated lineages, we took this SNV further for developing the diagnostic assay.

**Figure 1:**
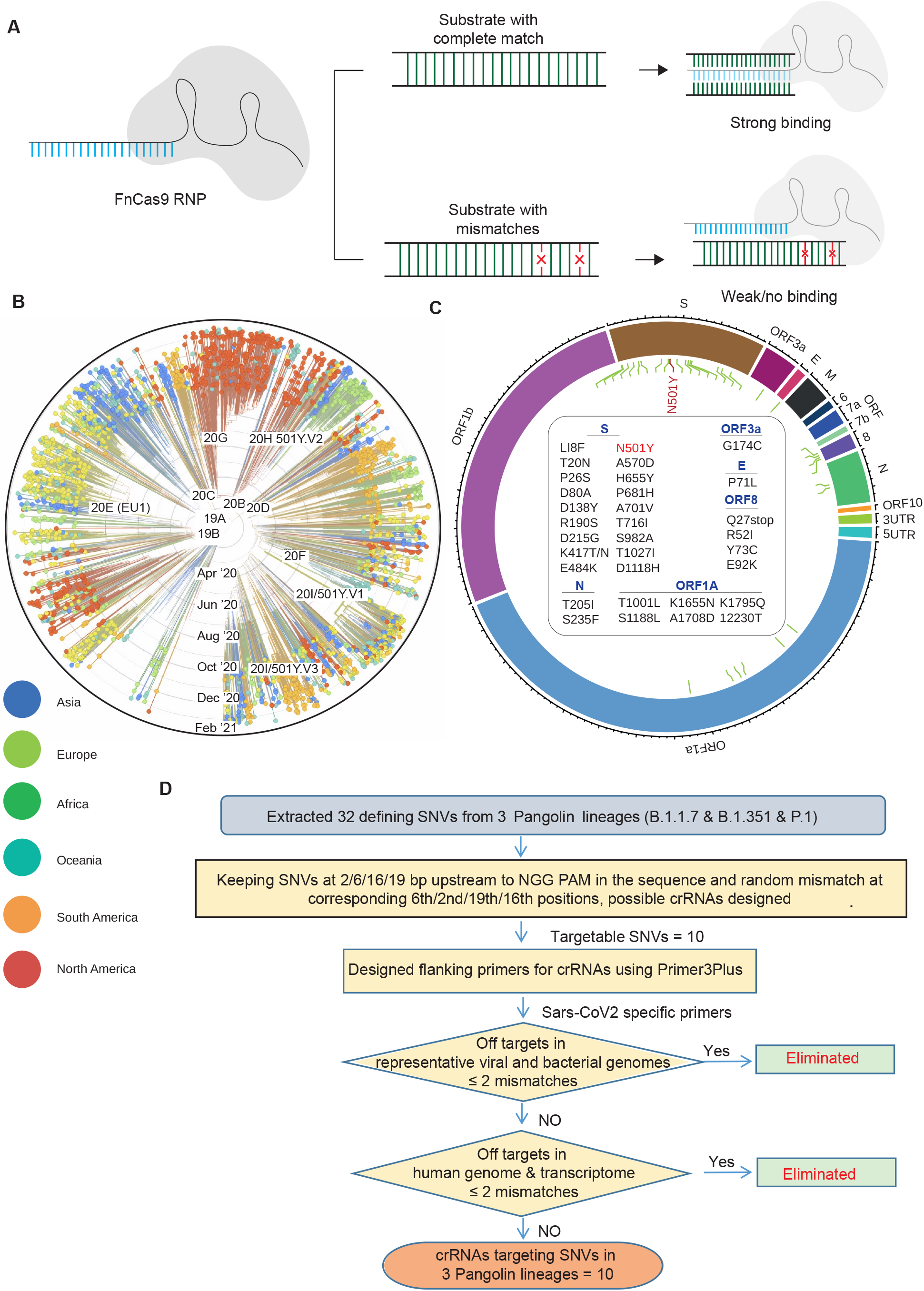
Schematic for RAY. **A**, FnCas9 is unable to bind or cleave targets having two mismatches at the PAM proximal 2nd and 6th positions as shown. **B**, Phylogeny plot from Nextstrain^39^ shows evolution of SARS-CoV2 viruses globally. Samples are plotted from the first reported case (September 15th) of new UK variant (20I/501Y.V1 or B.1.1.7). **C**, Circle plot showing the position of the different mutations in the genome of SARS-CoV2. N501Y(A23063T), which is common across all variants, is highlighted. **D**, A pipeline for the design criteria for RAY specific oligos and sgRNAs is shown.

We designed primer pairs surrounding the N501Y mutation after analyzing the mutational spectrum in SARS-CoV2 strains obtained from the publicly available sequencing data^5,37,43^. Additionally, we ensured that no regions from human or non-human transcriptome shared significant homology to the sites to prevent any non-specific amplification during the reverse transcription PCR (RT-PCR) reaction (Figure 1D). Our earlier FELUDA design validated on a large number of patient samples used an S-gene sgRNA for detecting the presence of SARS-CoV2 inpatient samples^25^. We reasoned that including the same sgRNA for detecting the S gene region which is in the vicinity of the N501Y variant would serve as an internal positive control for the presence of the SARS-CoV2 virus in the sample. To this end, we ensured that all amplicons contained both the sites of the N501Y targeting sgRNA as well as the earlier FELUDA S-gene sgRNA.

### RAY can successfully discriminate N501Y and WT nCoV2 substrates

We next tested the N501Y sgRNA containing the mismatch at PAM proximal 6th position to selectively bind and cleave the mutant substrate, while not affecting the WT substrate due to mismatch at PAM proximal 2nd and 6th positions. We found that FnCas9 was able to successfully cleave the substrate containing the N501Y mutation while the WT sequence was not cleaved (Figure 2A). Importantly, this leads to a distinct pattern on agarose gel that can distinguish the two variants. We established that this approach for variant identification can be performed using a stand-alone or portable electrophoresis apparatus leading to a turnaround time of about 1.5 h from RNA to read-out and can readily replace the current sequencing-based discrimination of variants for rapid diagnosis (Figure 2A).

**Figure 2:**
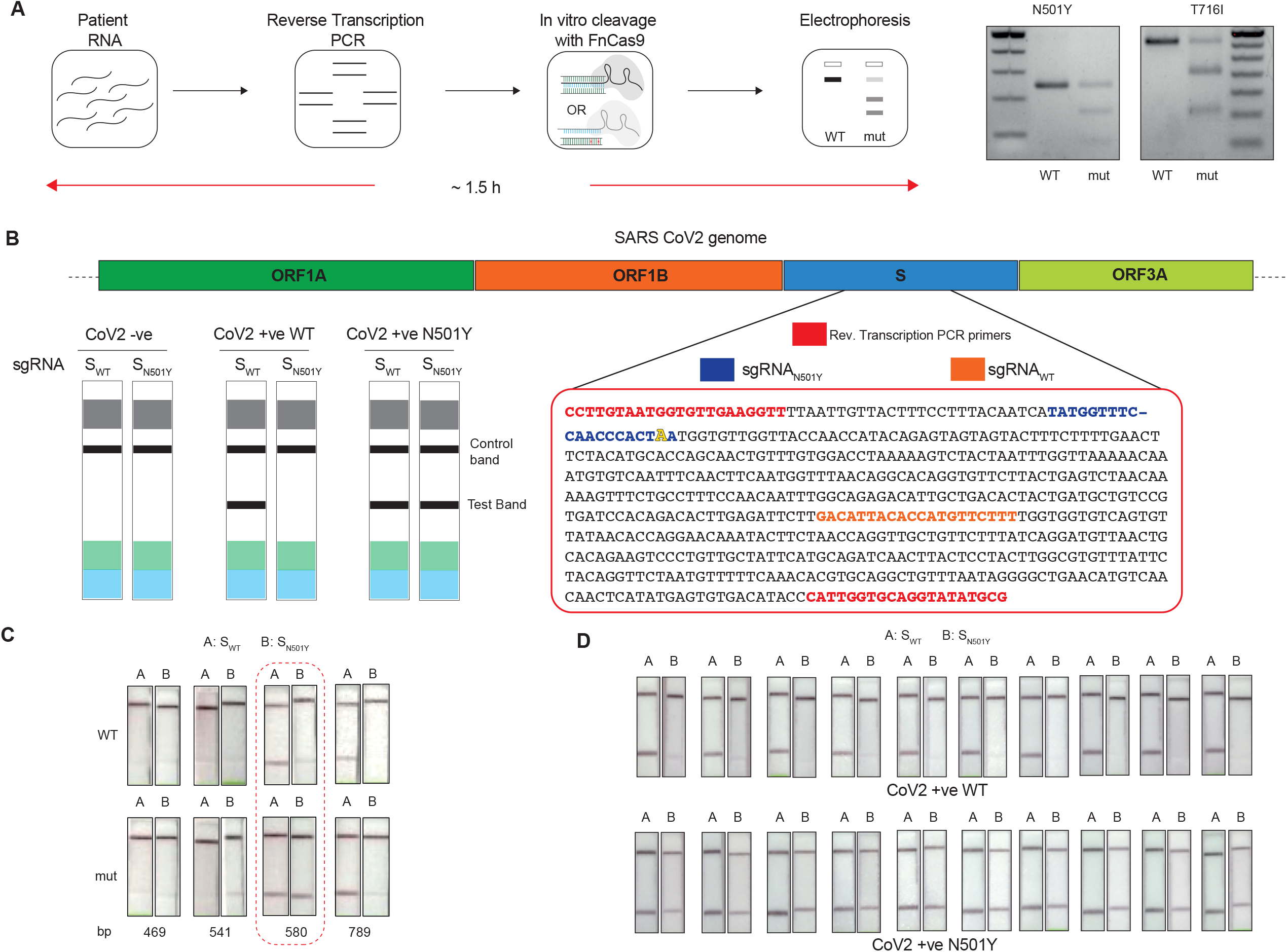
Adaptation of RAY for identification of N501Y mutation. **A**, Schematic showing the application of RAY for distinguishing variants (N501Y and T716I) using electrophoresis, key steps in the process are shown. **B**, Left panel showing the different outcomes of RAY on a paper strip based on the starting material. Distinct bands on the streptavidin line (test line) characterize CoV2 negative, CoV2 wild type and CoV2 N5012Y variants. Right panel shows the sequence, primers and sgRNAs (indicated) for the 580 bp amplicon. The N501Y mutation position is shown in raised font. **C**, RAY optimization with different PCR amplicons of S gene. Optimal amplicon length denoted by dotted rounded box. **D**, Reproducibility in output on multiple runs of RAY on the same samples (WT or N501Y) showing high concordance between assays (n=10).

Since the CoV2 genome has already undergone multiple mutations, and there could be more mutations of concern occurring in the recent future, we validated RAY on another mutation T716I using the same design principles. Remarkably, T716I was also successfully distinguished using RAY (Figure 2A). All together, RAY design parameters are consistent across different mutations, and the pipeline can be seamlessly adapted for novel mutations that might emerge in the nCoV2 genome.

### RAY on a lateral flow device generates rapid and accurate diagnostic readout for N501Y mutation in patient samples

The portable electrophoresis based identification of the N501Y(A23063T) mutation can be adapted for COVID-19 detection under laboratory conditions. However, dye-based systems have low sensitivity and will undergo limitations when viral load is not high. We reasoned that for RAY to detect and diagnose variants, we need to adapt the detection to a more sensitive readout. Since the FELUDA platform on a lateral flow strip can detect COVID-19 positive individuals across all ranges of Ct values, we developed a pipeline for adapting RAY on a paper strip.

To generate a visually distinctive signal between WT and mutant variants, we modified our previous FELUDA protocol to identify SNV accurately in the sample^25^. Firstly, we performed a single-step reverse transcription PCR to generate a biotin-labeled amplified product that can be detected by a single sgRNA (S-gene) if the sample is WT and by both sgRNAs (S-gene and N501Y) if the sample contains the N501Y variant. A single amplicon for both the sgRNAs ensures that there is no PCR bias in detecting the two regions by CRISPRDx from the same sample. Secondly, we eliminated one of the biotin label primers (from the FELUDA reaction) to reduce the chances of background signal due to non-specific interactions of the unused biotin primers with the streptavidin. Thirdly, we made modifications at the level of amplicon length, PCR conditions, and RNP concentration to generate the most optimal protocol that can produce a clear discernible signal on a commercially available paper strip (Figure 2B, Methods). To validate the reproducibility of this method, we took mutant and WT substrates, and performed RAY ten times, and were able to successfully distinguish them on every occasion (Figure 2C). Finally, we tested RAY on RNA extracted from samples of RT-PCR positive SARS CoV2 infected individuals who returned to India from the United Kingdom between December 2020 and January 2021. The samples were also sequenced to detect the presence of N501Y mutation (Oxford Nanopore platform). RAY, was able to correctly identify the variant signature in all eight samples that harbored the mutation (Figure 3A). Similarly, RAY correctly classified all eight samples where neither the N501Y mutation nor any other lineage variants were present. Besides, RAY identified all confirmed COVID-19 negative samples (twelve) tested simultaneously (Figure 3A). Importantly, RAY identified samples across all ranges of Ct values showing that the assay is significantly robust across a large range of viral titre. As with FELUDA, visual readout of RAY can be supplemented at higher Ct values by the smart-phone based application TOPSE to generate background-corrected intensity values. Notably, for every sample, the whole process from RNA to read-out can be completed in ∼75 min, highlighting the efficacy of assay for the quick identification of individuals carrying the variant (Figure 3B).

**Figure 3:**
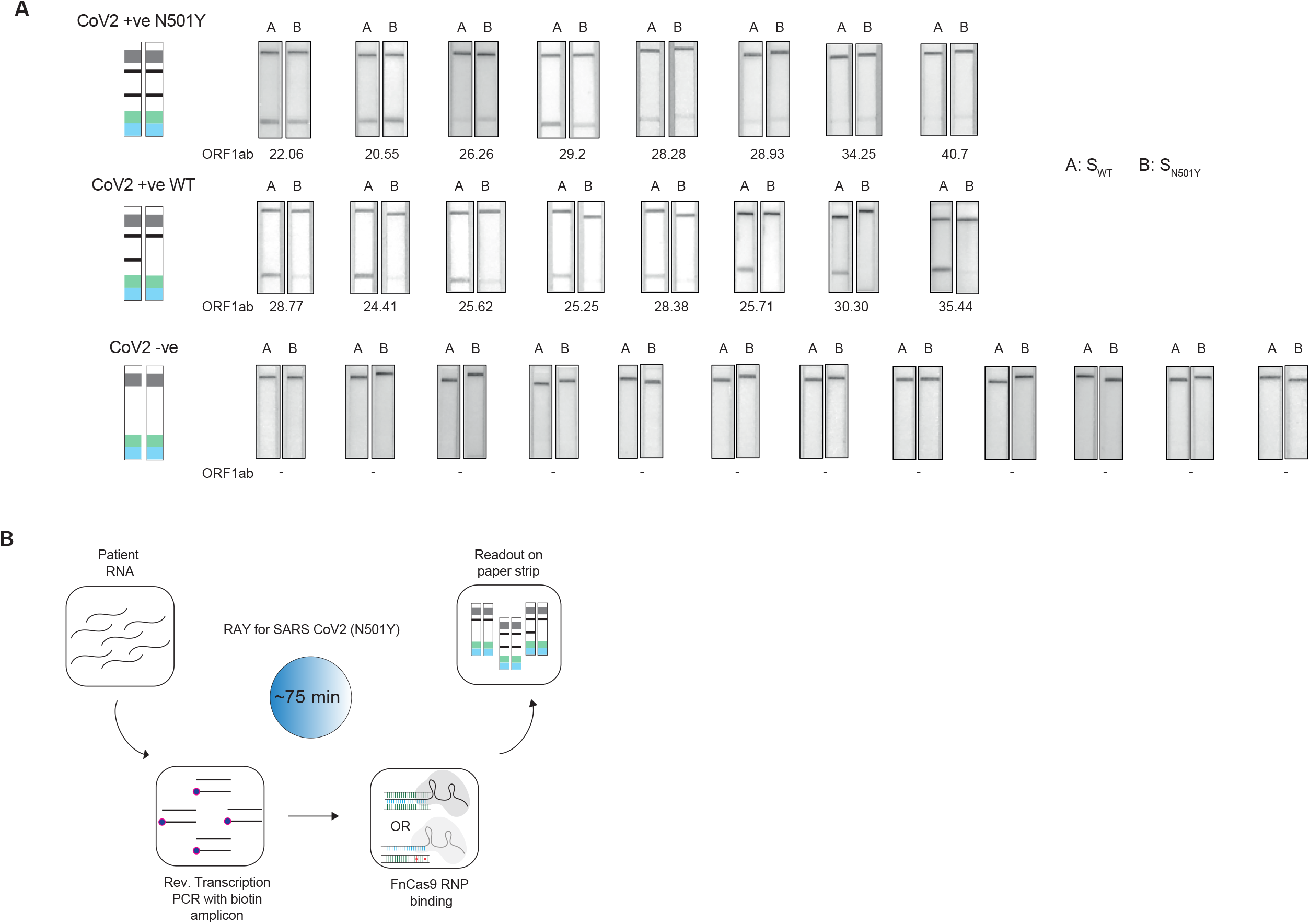
Validation of RAY on patient samples. **A**, RAY outcomes on three groups of patient samples as indicated. The ORF1 Ct values for every sample is indicated below. **B**, Schematic of the RAY assay showing individual steps in the pipeline.

## DISCUSSION

Currently, deep-sequencing of patient samples is routinely being followed by health authorities in various countries to track and isolate cases containing the CoV2 variants^21-24^. The accuracy of variant calling in any sequencing platform is dependent on the very high quality and coverage of reads to the reference genome. Since the process from sample to sequence consists of multiple steps, low viral titre can be missed out during sequencing assays reducing the overall sensitivity. The RAY pipeline uses an amplification step and enzymatic binding assay and is reproducible across all the viral titre. Therefore can detect samples across all ranges of Ct values.

The main advantage of sequencing COVID-19 samples is the generation of cumulative information about all mutations co-existing in a given variant^44,,45^. This information is valuable to track and trace how lineages evolve and has led to the identification of mutations in the first place. However, routine sequencing of patient samples for surveillance or First point detection is fraught with issues, such as cost, logistics, and turnaround times. A typical deep sequencing cost per sample is several orders of magnitude more than RAY (which costs less than 10 USD per sample) and the minimum turnaround time for sequencing is 36 hours (Supplementary Table 2). Importantly, sequencing runs are generally done on pooled samples to save on cost, workforce, and machine time, all of which necessitate the advent of alternate solutions that can identify variants through a simple pipeline. RAY,-on the contrary- is scalable to any number of samples at a given time.

RAY gives the advantage of rapid identification of a conserved mutation and is useful for diagnosis and surveillance. Since its readout is on a paper strip where one mutation can be detected at a time, identifying multiple mutations will require performing several RAY reactions. Additionally, RAY is not suited for identifying new variants that have emerged in a sample-unlike sequencing. However, the universality of the sgRNA design by which FnCas9 can discriminate between mutations makes it possible to multiplex the identification of several variants from the same sample.

Sequencing generates a large amount of data and requires physical storage and high-performance computing methodologies to record, analyze and interpret the data^44,45^. In addition, dedicated personnel with a significant understanding of sequence analysis and programming skills are necessary for identifying variants within reads generated from a sample. In contrast, RAY uses a visual readout on a paper strip leading to a binary decision about the sample. That has the advantage of rapid screening, particularly at the point of care settings. To facilitate electronic recording and analysis of the readout TOPSE, a mobile app described in our earlier study can be used in combination with RAY^25^. In addition to background correction, it also enables an automated decision-making process to assist the user’s visual interpretation^25^.

RAY has the scope of improvement in both performance and robustness. The use of chemically modified crRNAs would lead to greater stability of reaction components and higher band intensities, particularly for samples with high Ct values. Similarly, as shown earlier for FELUDA, a fluorescence-based readout can facilitate multiplexing several samples in a single run. This is useful for scaling up of testing, particularly where rapid decision making is required.

In this study, we report the development of a CRISPR diagnostic platform for detecting variants in SARS CoV2. Considering the rise and spread of mutated lineages of the virus in multiple countries, it is imperative to develop technologies that can quickly and accurately detect these variants. Since the first generation vaccines against SARS CoV2 have been developed against the parent strain, careful assessment of the variant lineages and their response to the vaccine will become imperative in the coming months of the pandemic^46-48^. Novel variants, particularly those correlated with adverse disease manifestation, need to be controlled so that these do not spread rampantly across populations. Testing and isolating them will thus continue to be at the forefront of disease control measures.

## MATERIALS AND METHODS

### Study Design

The study was designed to evaluate the efficacy of RAY on clinical samples. The intent of the study was to develop a robust CRISPR diagnostic that can perform with high accuracy for variant detection at significantly low cost and time taken for detection. For the N501Y detection using FELUDA, SARS CoV2 and control RNA samples were received from the diagnostic laboratory at CSIR-Institute of Genomics and Integrative Biology, where the clinical diagnosis was done before the left-over samples were used in this study.

### Oligos

A list of all oligos (Merck) used in the study can be found in Supplementary Table 3.

### Protein purification

Plasmids containing FnCas9-WT and dFnCas9 (Acharya, S. et al. Proc. Natl. Acad. Sci. 116, 20959–20968 (2019)) sequences were transformed and expressed in Escherichia coli Rosetta 2 (DE3) (Novagen). Rosetta 2 (DE3) cells were cultured at 37°C in LB medium (with 50mg/ml Kanamycin) and induced when OD600 reached 0.6, using 0.5mM isopropyl β-D-thiogalactopyranoside (IPTG). After an overnight culture at 18°C, E.coli cells were harvested by centrifugation and resuspended in a lysis buffer (20mM HEPES, pH 7.5, 500mM NaCl, 5% glycerol supplemented with 1X PIC (Roche) containing 100ug/ml lysozyme. Subsequent cell lysis by sonication, the lysate was put through to Ni-NTA beads (Roche). The eluted protein was further purified by size-exclusion chromatography on HiLoad Superdex 200 16/60 column (GE Healthcare) in a buffer solution with 20 mM HEPES pH 7.5, 150 mM KCl, 10% glycerol, 1mM DTT. The purified proteins were quantified by Pierce BCA protein assay kit (Thermo Fisher Scientific) and stored at -80°C until further use.

### RAY crRNA and primer design

All 32 mutations defining B.1.1.7 and B.1.351/501Y-V2 lineages were extracted from Pangolin^37,49^. Further by keeping these mutations at 2nd/6th/16th/19th bp upstream to PAM (NGG) in SARS-CoV2 we were able to target 10 mutations. Next, these mutations were incorporated into the crRNA sequence followed by a random mismatch at corresponding 6th/2nd/19th/16th position. Primers flanking these crRNAs were designed using Primer3plus python library^50^. Finally, Primers were investigated for off-targets on representative bacterial genome database (NCBI), virus genome database (NCBI) and human genome/transcriptome (GENCODE GRCh38)^51-53^.

### RAY Detection assays

Sequences containing WT and Mutant were reverse transcribed, and amplified using primers with/without 5’ biotinylation from SARS-CoV2 Viral RNA enriched samples.

### Detection *via In vitro* Cleavage (IVC)

Purified PCR amplicons containing WT and Mutant sequences were used as substrate in *in vitro* cleavage assays, as optimized in our previous studies. Substrate amplicons were treated with reconstituted RNP complex (100nM) in a reaction buffer (20 mM HEPES, pH7.5, 150mM KCl, 1mM DTT, 10% glycerol, 10mM MgCl2) at 37°C for 10 minutes, and cleaved products were visualized on a 2 % agarose gel.

### RAY *via* Lateral Flow Assay (RT-PCR)

A region from the SARS-CoV2 S gene containing N501Y/T716I mutations was reverse transcribed and amplified using a single end 5’ biotin-labeled primers.

*In vitro* transcription of sgRNAs/crRNAs was done using MegaScript T7 Transcription kit (ThermoFisher Scientific) following manufacturer’s protocol and purified by NucAway spin column (ThermoFisher Scientific). Chimeric gRNA (crRNA:TracrRNA) was prepared by equally (crRNA:TracrRNA molar ratio,1:1) combining respective crRNAs and synthetic 3’-FAM-labeled TracrRNA in an annealing buffer (100mM NaCl, 50mM Tris-HCl pH8 and 1mM MgCl2) by heating at 95°C for 2-5 minutes and then allowed to cool at room temperature for 15-20 min. RNP complex was prepared by equally mixing (Protein:sgRNA molar ratio,1:1) Chimeric gRNA and dead FnCas9 in a buffer (20mM HEPES, pH7.5, 150mM KCl, 1mM DTT, 10% glycerol, 10mM MgCl2) and rested for 10 min at RT. Target biotinylated amplicons were then treated with the RNP complexes for 10 min at 37°C. Finally, 80ul of Dipstick buffer was added to the mix along with Milenia HybriDetect 1 lateral flow strip. The strips were allowed to stand in the solution for 2-5 minutes at room temperature and the result was observed. The strip images can be processed using TOPSE application that generates background corrected values from the smart phone acquired images of the strip. This application has been previously trained on a large number of CoV2 samples^25^.

### qRT-PCR SARS-CoV-2 detection

qRT-PCR was performed using STANDARD M nCoV Real-Time Detection kit (SD Biosensor) as per manufacturer’s protocol. Briefly, per reaction 3µl of RTase mix and 0.25µl of Internal Control A was added to 7µl of the reaction solution. 5µl of each of the negative control, positive control, and patient sample nucleic acid extract was added to the PCR mixture dispensed in each reaction tube. The cycling conditions on the instrument were as follows: Reverse transcription 50°C for 15 minutes, Initial denaturation 95°C for 1 min, 5 Pre-amplification cycles of 95°C for 5 sec; 60°C for 40 sec followed by 40 amplification cycles of 95°C for 5 sec; 60°C for 40 sec. Signal was captured in the FAM channel for the qualitative detection of the new coronavirus (SARS-CoV-2) ORF1ab (RdRp) gene, JOE (VIC or HEX) channel for E gene, and CY5 channel for internal reference.

### Sequencing of patient samples

The individuals with UK travel history between December 2020-January 2021, who were found to be RT-PCR positive for SARS-CoV2, were sequenced to determine whether it was a UK variant (20I/501Y.V1) or non-UK variant. With a low number of samples arriving each day in the lab and with quick turnaround time for reporting, Nanopore sequencing was undertaken.

### SARS-CoV-2 Whole genome sequencing using Nanopore platform

In brief, 100ng total RNA was used for double-stranded cDNA synthesis by using Superscript IV (ThermoFisher Scientific, Cat.No. 18091050) for first strand cDNA synthesis followed by RNase H digestion of ssRNA and second strand synthesis by DNA polymerase-I large (Klenow) fragment (New England Biolabs, Cat. No. M0210S). Double stranded cDNA thus obtained was purified using AMPure XP beads (Beckman Coulter, Cat. No. A63881). The SARS-CoV-2 genome was then amplified from 100ng of the purified cDNA following the ARTIC V3 primer protocol. Sequencing library preparation consisting of End Repair/dA tailing, Native Barcode Ligation, and Adapter Ligation was performed with 200ng of the multiplexed PCR amplicons according to Oxford Nanopore Technology (ONT) library preparation protocol-PCR tiling of COVID-19 virus (Version: PTC_9096_v109revE_06Feb2020). Sequencing in sets of 24 barcoded samples was performed on MinION Mk1B platform by ONT.

### Nanopore Sequencing analysis

The ARTIC end-to-end pipeline was used for the analysis of MinION raw fast5 files up to the variant calling. Raw fast5 files of samples were basecalled and demultiplexed using Guppy basecaller that uses the base calling algorithms of Oxford Nanopore Technologies (https://community.nanoporetech.com) with phred quality cut-off score >7 on GPU-linux accelerated computing machine. Reads having phred quality scores less than 7 were discarded to filter the low-quality reads. The resulting demultiplexed fastq were normalized by read length of 300-500 (approximate size of amplicons) for further downstream analysis and aligned to the SARS-CoV-2 reference (MN908947.3) using the aligner Minimap2 v2.17^54^. Nanopolish^55^ were used to index raw fast5 files for variant calling from the minimap output files. To create consensus fasta, bcftools v1.8 was used over normalized minimap2 output. Sample sequences have been uploaded to GISAID repository.

## Data Availability

All data presented in the manuscript will be made available upon reasonable request to the corresponding authors.

## Acknowledgments

We thank all members of Chakraborty and Maiti labs for helpful discussions and valuable insights. This study was funded by CSIR Sickle Cell Anemia Mission (HCP0008), TATA Steel CSR (SSP2001) and a Lady Tata Young Investigator award (GAP0198) to D.C. The sequencing was funded by the CSIR (MLP-2005), Fondation Botnar (CLP-0031), Intel (CLP-0034) and IUSSTF (CLP-0033). We also acknowledge Dr. Mohd. Faruq, Shaista khan and Sarfaraz Alam (CSIR IGIB) for help with isolation of RNA from the VTM of the RT-PCR positive samples.

## Contributions

D.C., M.K., S.G. and S.M. conceived, designed and interpreted the experiments. A.H.A provided bioinformatics support.. Rh.P., S.A. and P.K. contributed to studies for single mismatch discrimination using RAY. A.K., R.M., J.S.V., A.M. and Ra.P. contributed to sequencing and analysis of the COVID-19 samples. D.C. wrote the manuscript with inputs from M.K. and S.G. and S.M.

## Ethics declarations

The present study was approved by the Ethics Committee, Institute of Genomics and Integrative Biology, New Delhi. A patent application has been filed in relation to this work.

## Supplementary Tables

**Table 1**: Mutations across CoV2 lineages showing the possibility of FnCas9 based targeting based on RAY

**Table 2**: Price estimate of RAY consumables per reaction

**Table 3**: List of oligos used in this study

## References

1. Rabi, Firas A., et al. “SARS-CoV-2 and coronavirus disease 2019: what we know so far.” Pathogens 9.3 (2020): 231.

2. Pachetti, Maria, et al. “Emerging SARS-CoV-2 mutation hot spots include a novel RNA-dependent-RNA polymerase variant.” Journal of translational medicine 18 (2020): 1–9.

3. Mercatelli, Daniele, and Federico M. Giorgi. “Geographic and genomic distribution of SARS-CoV-2 mutations.” Frontiers in microbiology 11 (2020): 1800.

4. Chand, M. “Investigation of novel SARS-COV-2 variant: Variant of Concern 202012/01 (PDF).” Public Health England. PHE (2020).

5. https://www.cogconsortium.uk/news_item/the-new-sars-cov-2-variant-and-other-mutations-under-review-by-cog-uk/

6. NERVTAG paper on COVID-19 variant of concern B.1.1.7: paper from the New and Emerging Respiratory Virus Threats Advisory Group (NERVTAG) on new coronavirus (COVID-19) variant B.1.1.7. https://www.gov.uk/government/publications/nervtag-paper-on-covid-19-variant-of-concern-b117

7. du Plessis, Louis, et al. “Establishment and lineage dynamics of the SARS-CoV-2 epidemic in the UK.” Science (2021).

8. Tegally, Houriiyah, et al. Emergence and rapid spread of a new severe acute respiratory syndrome-related coronavirus 2 (SARS-CoV-2) lineage with multiple spike mutations in South Africa.” medRxiv (2020).

9. Resende, Paola Cristina, et al. “Spike E484K mutation in the first SARS-CoV-2 reinfection case confirmed in Brazil, 2020.”

10. Faria, N. R., et al. “Genomic characterisation of an emergent SARS-CoV-2 lineage in Manaus: preliminary findings.” (2021).

11. Roser, Max, et al. “Coronavirus pandemic (COVID-19).” Our world in data (2020).

12. Mackay, Ian M., Katherine E. Arden, and Andreas Nitsche. “Real-time PCR in virology.” Nucleic acids research 30.6 (2002): 1292–1305.

13. Carter, Linda J., et al. “Assay techniques and test development for COVID-19 diagnosis.” (2020): 591–605.

14. Emery, Shannon L., et al. “Real-time reverse transcription–polymerase chain reaction assay for SARS-associated coronavirus.” Emerging infectious diseases 10.2 (2004): 311.

15. Dramé, Moustapha, et al. “Should RT-PCR be considered a gold standard in the diagnosis of Covid-19?.” Journal of medical virology 92.11 (2020): 2312–2313.

16. Khan, Kashif Aziz, and Peter Cheung. “Presence of mismatches between diagnostic PCR assays and coronavirus SARS-CoV-2 genome.” Royal Society open science 7.6 (2020): 200636.

17. Afzal, Adeel. “Molecular diagnostic technologies for COVID-19: Limitations and challenges.” Journal of advanced research (2020).

18. Tahamtan, Alireza, and Abdollah Ardebili. “Real-time RT-PCR in COVID-19 detection: issues affecting the results.” Expert review of molecular diagnostics 20.5 (2020): 453–454.

19. Little, Stephen. “Amplification-refractory mutation system (ARMS) analysis of point mutations.” Current protocols in human genetics 7.1 (1995): 9–8.

20. Ahlawat, Sonika, et al. “Designing, optimization and validation of tetra-primer ARMS PCR protocol for genotyping mutations in caprine Fec genes.” Meta gene 2 (2014): 439–449.

21. Vandenberg, Olivier, et al. “Considerations for diagnostic COVID-19 tests.” Nature Reviews Microbiology (2020): 1–13.

22. McNamara, Ryan P., et al. “High-density amplicon sequencing identifies community spread and ongoing evolution of SARS-CoV-2 in the Southern United States.” Cell reports 33.5 (2020): 108352.

23. Xiao, Minfeng, et al. “Multiple approaches for massively parallel sequencing of SARS-CoV-2 genomes directly from clinical samples.” Genome medicine 12.1 (2020): 1–15.

24. Bhoyar, Rahul C., et al. “High throughput detection and genetic epidemiology of SARS-CoV-2 using COVIDSeq next generation sequencing.” bioRxiv (2020).

25. Azhar, Mohd, et al. “Rapid, accurate, nucleobase detection using FnCas9.” medRxiv (2020).

26. Patchsung, Maturada, et al. “Clinical validation of a Cas13-based assay for the detection of SARS-CoV-2 RNA.” Nature Biomedical Engineering 4.12 (2020): 1140–1149.

27. Ding, Xiong, et al. “Ultrasensitive and visual detection of SARS-CoV-2 using all-in-one dual CRISPR-Cas12a assay.” Nature communications 11.1 (2020): 1–10.

28. Lucia, Curti, Pereyra-Bonnet Federico, and Gimenez Carla Alejandra. “An ultrasensitive, rapid, and portable coronavirus SARS-CoV-2 sequence detection method based on CRISPR-Cas12.” bioRxiv (2020).

29. Broughton, James P.,et al. “CRISPR–Cas12-based detection of SARS-CoV-2.” Nature Biotechnology (2020): 1–5.

30. Wang, Rui, et al. “opvCRISPR: One-pot visual RT-LAMP-CRISPR platform for SARS-cov-2 detection.” Biosensors and Bioelectronics 172 (2020): 112766.

31. Guo, Lu, et al. “SARS-CoV-2 detection with CRISPR diagnostics.” Cell Discovery 6.1 (2020): 1–4.

32. Joung, Julia, et al. “Detection of SARS-CoV-2 with SHERLOCK one-pot testing.” New England Journal of Medicine 383.15 (2020): 1492–1494.

33. Fozouni, Parinaz, et al. “Amplification-free detection of SARS-CoV-2 with CRISPR-Cas13a and mobile phone microscopy.” Cell (2020).

34. Brogan, Daniel J., et al. “A Sensitive, Rapid, and Portable CasRx-based Diagnostic Assay for SARS-CoV2.” medRxiv (2020).

35. Santiago-Frangos, Andrew, et al. “Intrinsic Signal Amplification by Type-III CRISPR-Cas Systems Provides a Sequence-Specific Viral Diagnostic.” medRxiv (2020).

36. Yoshimi, Kazuto, et al. “Rapid and accurate detection of novel coronavirus SARS-CoV-2 using CRISPR-Cas3.” medRxiv (2020).

37. https://cov-lineages.org/global_report.html

38. Acharya, Sundaram, et al. “Francisella novicida Cas9 interrogates genomic DNA with very high specificity and can be used for mammalian genome editing.” Proceedings of the National Academy of Sciences 116.42 (2019): 20959–20968.

39. Hadfield, James, et al. “Nextstrain: real-time tracking of pathogen evolution.” Bioinformatics 34.23 (2018): 4121–4123.

40. Yi, Chunyan, et al. “Key residues of the receptor binding motif in the spike protein of SARS-CoV-2 that interact with ACE2 and neutralizing antibodies.” Cellular & molecular immunology 17.6 (2020): 621–630.

41. Zahradnik, Jiri, et al. “SARS-CoV-2 RBD in vitro evolution follows contagious mutation spread, yet generates an able infection inhibitor.” bioRxiv (2021).

42. Gu, Hongjing, et al. “Adaptation of SARS-CoV-2 in BALB/c mice for testing vaccine efficacy.” Science 369.6511 (2020): 1603–1607.

43. Shu, Yuelong, and John McCauley. “GISAID: Global initiative on sharing all influenza data–from vision to reality.” Eurosurveillance 22.13 (2017): 30494.

44. Viehweger, Adrian, et al. “Direct RNA nanopore sequencing of full-length coronavirus genomes provides novel insights into structural variants and enables modification analysis.” Genome research 29.9 (2019): 1545–1554.

45. Bull, Rowena A., et al. “Analytical validity of nanopore sequencing for rapid SARS-CoV-2 genome analysis.” Nature communications 11.1 (2020): 1–8.

46. Dong, Yetian, et al. “A systematic review of SARS-CoV-2 vaccine candidates.” Signal transduction and targeted therapy 5.1 (2020): 1–14.

47. Zhang, Yanjun, et al. “Safety, tolerability, and immunogenicity of an inactivated SARS-CoV-2 vaccine in healthy adults aged 18–59 years: a randomised, doubleblind, placebo-controlled, phase 1/2 clinical trial.” The Lancet Infectious Diseases (2020).

48. Hodgson, Susanne H., et al. “What defines an efficacious COVID-19 vaccine? A review of the challenges assessing the clinical efficacy of vaccines against SARS-CoV-2.” The lancet infectious diseases (2020).

49. Rambaut, Andrew, et al. “A dynamic nomenclature proposal for SARS-CoV-2 lineages to assist genomic epidemiology.” Nature microbiology 5.11 (2020): 1403–1407.

50. Untergasser, Andreas, et al. “Primer3—new capabilities and interfaces.” Nucleic acids research 40.15 (2012): e115–e115.

51. Brister, J. R., et al. “NCBI viral Genomes resource. Nucleic 553 Acids Res. 43.” D571–D577 554 (2015).

52. “Database resources of the national center for biotechnology information.” Nucleic acids research 46, no. D1 (2018): D8–D13.

53. Frankish, Adam, et al. “GENCODE reference annotation for the human and mouse genomes.” Nucleic acids research 47.D1 (2019): D766–D773.

54. Li, Heng. “Minimap2: pairwise alignment for nucleotide sequences.” Bioinformatics 34.18 (2018): 3094–3100.

55. Loman, Nicholas J., Joshua Quick, and Jared T. Simpson. “A complete bacterial genome assembled de novo using only nanopore sequencing data.” Nature methods 12.8 (2015): 733–735.

